# A novel open access multimodal dataset of nodule imaging and circulating proteome from a lung cancer screening cohort

**DOI:** 10.64898/2025.12.23.25342921

**Authors:** Miriam Cobo, Diego Serrano, Jennifer Barranco, Andrea Pasquier, Juan Pablo de-Torres, Javier J Zulueta, José Ignacio Echeveste, Ana Ezponda, Jesús Pueyo, Allan Argueta, Julián Sanz-Ortega, Juan Bertó, Ana Belén Alcaide, Madeleine Di Frisco, Carmen Felgueroso, Arantza Campo, Alejandra de la Fuente, Ana Escobar, Karmele Valencia, Daniel Orive, Maria del Mar Ocón, Hanna Beata Globacka, Maria Antonia Fortuño, Valerio Perna, María Rodríguez, María Dolores Lozano, Alfonso Calvo, Ruben Pio, Rayjean J. Hung, Luis M Seijo, Wilson Silva, Gorka Bastarrika, Lara Lloret Iglesias, Luis M. Montuenga

**Author notes:** Both authors share Senior authorship (Lara Lloret Iglesias, Luis M. Montuenga). Corresponding author: Luis M Montuenga.

## Abstract

**Introduction:** Low-dose computed tomography (LDCT) lung cancer screening has significantly enhanced early detection and patient survival rates in the population at risk. Current screening methods, that primarily rely on LDCT imaging, will very likely benefit from molecular biomarkers to achieve a more comprehensive, accurate, personalized and non-invasive risk assessment leveraging multimodal tools. We present a novel open access multimodal (imaging, proteomics and demographic) dataset designed to provide an available research resource on LDCT-based early lung cancer detection. The dataset includes annotated screening LDCT scans and plasma proteomics generated by proximity extension assay (Olink) platform.

**Methods:** The dataset integrates data from control screened individuals without nodules or with benign nodules, and LDCT-diagnosed lung cancer individuals, matched by sex, age and time between image and sample collection. Both radiological and molecular signatures were collected within a six month window, providing detailed insights into disease progression. Nodules were considered as lung cancer cases if biopsy-confirmed lung cancer was diagnosed within 5 years after imaging, enabling the study of longitudinal biomarker evolution and its correlation with imaging findings. To complement the dataset, clinical and demographic data are also available in open access, providing a detailed overview of patient characteristics. The informed consent signed by the participants allows for unrestricted open access for requests directy or indirectly related to lung cancer research.

**Results:** The dataset consists of annotated screening LDCT scans and plasma proteomics data measured with most of the Olink Target 96 platforms (1078 individual proteins across 12 panels focused on a specific area of disease or biology) for a total of 211 screening participants. There are 67 lung cancer patients, 68 matched controls with benign pulmonary nodules, 71 matched controls without nodules and 5 surgically excised false positive lesions. Experiments were performed to assess the technical quality and provide a proof-of-concept of usability of the dataset, showing the alignment with findings from previous published studies.

**Conclusion:** This comprehensive dataset aims to facilitate research towards the development of personalized multimodal artificial intelligence models. We also aim to support the investigation of the relationship between imaging and molecular data, paving the way for more accurate understanding of early lung cancer biology. Finally, our open access dataset may help to develop or validate individualized risk prediction models that could significantly advance early lung cancer detection and intervention strategies.

## 1 Introduction

Lung cancer is a leading cause of cancer death worldwide [1, 2], and the second most commonly diagnosed cancer in both men and women in the United States in 2023 [3]. Low dose computed tomography (LDCT) screening has improved early diagnosis of lung cancer, decreasing lung cancer mortality among population at high risk [4–6]. The population at risk is defined by different risk models, all of which include age, smoking exposure and other clinical or demographic traits. The United States Centers for Medicare & Medicaid Services have defined their eligibility criteria for LDCT-based lung cancer screening for beneficiaries who are 50 to 77 years old, asymptomatic, current smokers or those who quit within the past 15 years, and have a smoking history of at least 20 pack-years [7]. Other models include additional risk components, or different ranges of age and smoking to further refine inclusion criteria [8]. One of the unmet clinical needs in lung cancer screening is assessing and managing indeterminate risk pulmonary abnormalities (nodules) found in the CT images. These indeterminate risk (neither clearly benign, nor highly suspicious of malignancy) pulmonary nodules (IPNs) pose a challenge, since they are found in approximately 20% − 40% of screening participants [9]. A similar clinical situation may happen outside screening, when incidental lung nodules are found when performing CT-based diagnostic studies for other conditions [10].

The Lung CT Screening Reporting and Data System (Lung-RADS) was developed to standardize the reporting and management of screen-detected pulmonary nodules, and improve risk assessment [11]. LungRADS classifies lung nodules on a scale from 1 to 4X, where 1-2 indicate benign nodules with minimal risk, 3 represents indeterminate nodules requiring follow-up, and 4A-4X suggest a high suspicion of malignancy, warranting further diagnostic evaluation. Highly suspicious lung nodules typically require a biopsy for definitive diagnosis, usually through an invasive procedure. Thus, precise evaluation of risk in pulmonary nodules found in the screening process (and also in the clinical finding of incidental nodules) is crucial to improve patient survival and avoid unnecessary biopsy procedures. Comprehensive evaluation after an IPN finding requires a close follow-up at different time points, in order to identify differential characteristics and progression patterns that enable monitoring risk and eventually early detection of a malignant lesion. Radiologist visual assessment of LDCT screening scans remains the gold standard approach for evaluating lung nodules in the clinics. This evaluation is labor-intensive, time-consuming and has some degree of subjectivity; expertise variability and subtle pixel-level differences in malignant nodules make accurate assessment a challenging task, and compromises the accuracy of this approach. Efforts have been made to standardize the quantitative lung nodule CT measurements [12, 13]. Computer-aided diagnosis (CAD) systems, particularly those leveraging deep learning (DL) techniques, have demonstrated strong potential for evaluating pulmonary nodule malignancy, providing valuable support to radiologists and improving the management of IPNs [14–16].

The majority of lung nodule CAD risk assessment DL based models in the literature rely on the use of the publicly available LIDC-IDRI dataset [17], which was collected by the US National Cancer Institute in 2011, as training tool. This database includes nodule malignancy annotations (ranging from label 1, indicating low risk, to label 5, corresponding to high risk) provided by four independent radiologists. However, this dataset lacks pathological examination gold standard labels, particularly for nodules rated as highly suspicious of malignancy (labels 4 and 5). There are few published publicly available datasets that contain gold standard labels for lung cancer nodules. The most comprehensive in terms of data volume is the National Lung Screening Trial (NLST) [18], a randomized controlled clinical trial of screening tests for lung cancer, where a subset of 28 000 LDCT images can be granted access through the Cancer Data Access System, including 623 participants with screen-detected cancer. However, nodule annotations are not available, although some studies have obtained them for subsets of the dataset [19, 20]. LUNGx challenge dataset [21] released a total of 83 scans in 2014, but all except 13 were contrast enhanced, which is not compatible with the usual lung cancer screening protocol. LIDP dataset [22] was claimed to be released after an embargo period. Yet, to the best of our knowledge, it is still not publicly available. Recently, two other CT datasets were published: one specifically annotated with histopathology-based information [23], and the other one was a cross spatio-temporal lung nodule dataset with pathological information for nodule identification [24]. These two latter datasets incorporating pathological gold standard labels constitute a step forward to improving CAD systems for lung cancer detection, but they were not obtained in the context of LDCT lung cancer screening. Both datasets were collected in Chinese cohorts, where existing guidelines for nodule management developed on US [4] and European [5] cohorts have shown suboptimal performance due to variations in the incidence of lung cancer, specifically in what relates to non- or low dose-smoking sensitivity, among Asian populations, particularly within the Chinese population [14, 25]. Hence, to ensure broad generalizability to other populations [26], further evaluation across diverse cohorts is essential, highlighting the need to expand publicly available datasets to support the scientific community in this field. This need is specially urgent in the context of LDCT-based lung cancer screening, where the development of artificial intelligence (AI) driven image analysis algorithms is accelerating, and relies on broad access to robust training datasets.

Pathological examination is the gold standard to confirm the presence of tumor in highly suspicious nodules. In contrast, benign nodules are not routinely biopsied, as the procedure is invasive, carries a risk of complications, and is generally reserved for cases where malignancy is suspected. As a result, there is no pathological confirmation ensuring the absence of malignancy in nodules classified as benign. Instead, their classification is based on long-term (at least two years) follow-up and nodule image stability over time. This follow-up process provides a temporary evaluation of nodules, allowing for the identification of those that remain stable and can be classified as benign, while highly suspicious cases are ultimately referred for biopsy. Thus, in radiological clinical practice, nodules can be categorized as highly suspicious, indeterminate (some of which may be considered suspicious in subsequent follow-ups), and low-risk (those that remain stable over time). In a few cases, biopsy excludes lung cancer in nodules initially deemed suspicious, revealing them as imaging false positives. Limiting dataset annotations to only those confirmed by histopathology, as in the dataset of Jian *et al*. [23, 24], reduces the effectiveness of CAD systems, since they are trained only on nodules that were already flagged as highly suspicious and referred to biopsy by the radiologist. This approach hinders the clinical usefulness, since in real-world scenarios there is a high proportion of IPNs, and they should be also used to train CAD systems. The absence of gold standard for highly suspicious nodules, as seen in LIDC-IDRI [17], means there is no definitive pathological confirmation, therefore, there exists the possibility of misdiagnosis [22]. This lack of gold standard introduces noise and uncertainty into CAD systems, leading to potential increases in false positives and false negatives.

Biomarkers have been proposed as potential adjuncts for the characterization of risk to add information to risk models to refine inclusion criteria, and as tools to help in the characterization of the potential malignancy of IPNs [27, 28]. Gene expression–based strategies have revolutionized breast cancer care by guiding treatment decisions with greater precision [29, 30]. Similarly, current lung cancer screening research is exploring circulating biomarkers in blood as non-invasive tools for diagnosis and risk estimation. These biomarker approaches, and specifically the analysis of circulating proteomics, aim to bring a higher degree of personalized care to lung cancer in line with genomics strategies in breast cancer. The Integrative Analysis of Cancer Risk and Etiology (INTEGRAL) research consortium program

[31] comprises 3 projects, among them the Risk Biomarker project aims to investigate novel prediagnostic circulating protein biomarkers to enhance lung cancer risk estimation [32]. The INTEGRAL Nodule Malignancy Project objectives are, among other, to identify circulating proteins that can differentiate benign versus malignant pulmonary nodules following an initial LDCT scan. The identification and validation of protein biomarkers provides additional information on lung cancer risk, and is a promising direction towards improving current image-based risk prediction models [31]. The present study describes the data included in the LDCT Pamplona-ELCAP screening cohort (P-ELCAP), collected at our institution, as one of the four screening cohorts included in the Nodule Malignancy project in the INTEGRAL program. The circulating proteome in prediagnostic plasma samples collected during LDCT screening was quantified with Olink proteomics platform using the proximity extension assay (PEA), as described in previous work [31, 33].

In this work, we aim to develop and publicly release an open-access dataset, including imaging and proteomics information, of this P-ELCAP novel lung cancer screening cohort for which multimodal data have been collected. The dataset includes the annotated LDCT image, the relative concentration of more than 1000 blood plasma circulating proteins markers, as well as several clinical and demographic variables. The dataset comprises data from 211 P-ELCAP LDCT-based screening participants, including 67 lung cancer cases, 68 matched controls with benign pulmonary nodules confirmed in subsequent follow-ups, and 71 matched controls without lung nodules. Moreover, there are 5 imaging false positives. Imaging and blood collection took place within a similar time range of *±* 6 months, always before surgery in the case of cancer cases. In the case of these biopsy confirmed tumors, the image and blood were collected within 5 years prior to diagnosis. Individuals selected for both control groups (with benign nodules and without nodules) were matched with cancer cases for age, sex and time of sample/image collection. In total, there are 138 nodules segmented in the dataset, 65 malignant (a few were not visible at the LDCT corresponding to the blood collection), 68 benign and 5 corresponding to imaging false positives. The dataset has been fully anonymized to prevent exposure of sensitive patient information. As aforementioned, this study is part of the P-ELCAP cohort collected at Clínica Universidad de Navarra (Spain) since 2000. To the best of our knowledge, this is the first pulmonary nodule dataset to incorporate annotated LDCT images together with molecular protein biomarkers to leverage multimodal early lung cancer diagnosis methods. We further demonstrate, through a proof-of-concept analysis, the potential of this dataset for personalized lung cancer risk assessment with a technical validation on LDCT image models, and Olink circulating protein markers.

The contributions and innovative aspects of this article can be summarized as follows:

- We have curated and provided open access to a novel multimodal early lung cancer screening dataset. The dataset incorporates annotated screening LDCT scans and plasma proteomics data measured with the Olink Target 96 platform (*>*1000 proteins) for a total of 211 screening participants.
- For lung cancer cases, pathological gold standard label confirmed the presence and subtype of tumor. There are 67 lung cancer patients (38 adenocarcinoma, 12 squamous cell carcinoma, 9 large cell carcinoma, 4 small cell carcinoma, 2 mixed adeno-squamous cell carcinoma and 2 other/NOS), 68 controls with benign pulmonary nodules, 71 controls without nodules and 5 surgically excised false positive lesions.
- We have conducted several experiments using existing classification models on the developed dataset. The results highlight inherent challenges associated with the limited dataset size. Nevertheless, due to its high quality and careful curation, the dataset holds significant value for external validation in other cohorts. Consequently, this dataset is highly valuable and represents an important step toward advancing the field of personalized medicine through AI-based systems in the context of LDCT-based lung cancer screening.

## 2 Materials and methods

The development and open access release of P-ELCAP dataset consisted of four steps: lung cancer screening sub-cohort selection of three matched groups of participants plus false positives; data acquisition and anonymization; data selection, curation and protein analysis; data annotation, preprocessing and dataset submission to the repository. In addition, two other steps are ongoing in our work to develop a multimodal nodule risk malignancy tool: implementation of baseline machine learning (ML) models, and final development of a fusion module ready for validation. The fusion model strategy integrating radiological and molecular signatures is a highly promising objective which is currently being carried out, and will be explored and validated in the next future. The six steps of the development of the multimodal nodule risk scoring tool are depicted in Figure 1.

**Figure 1.**
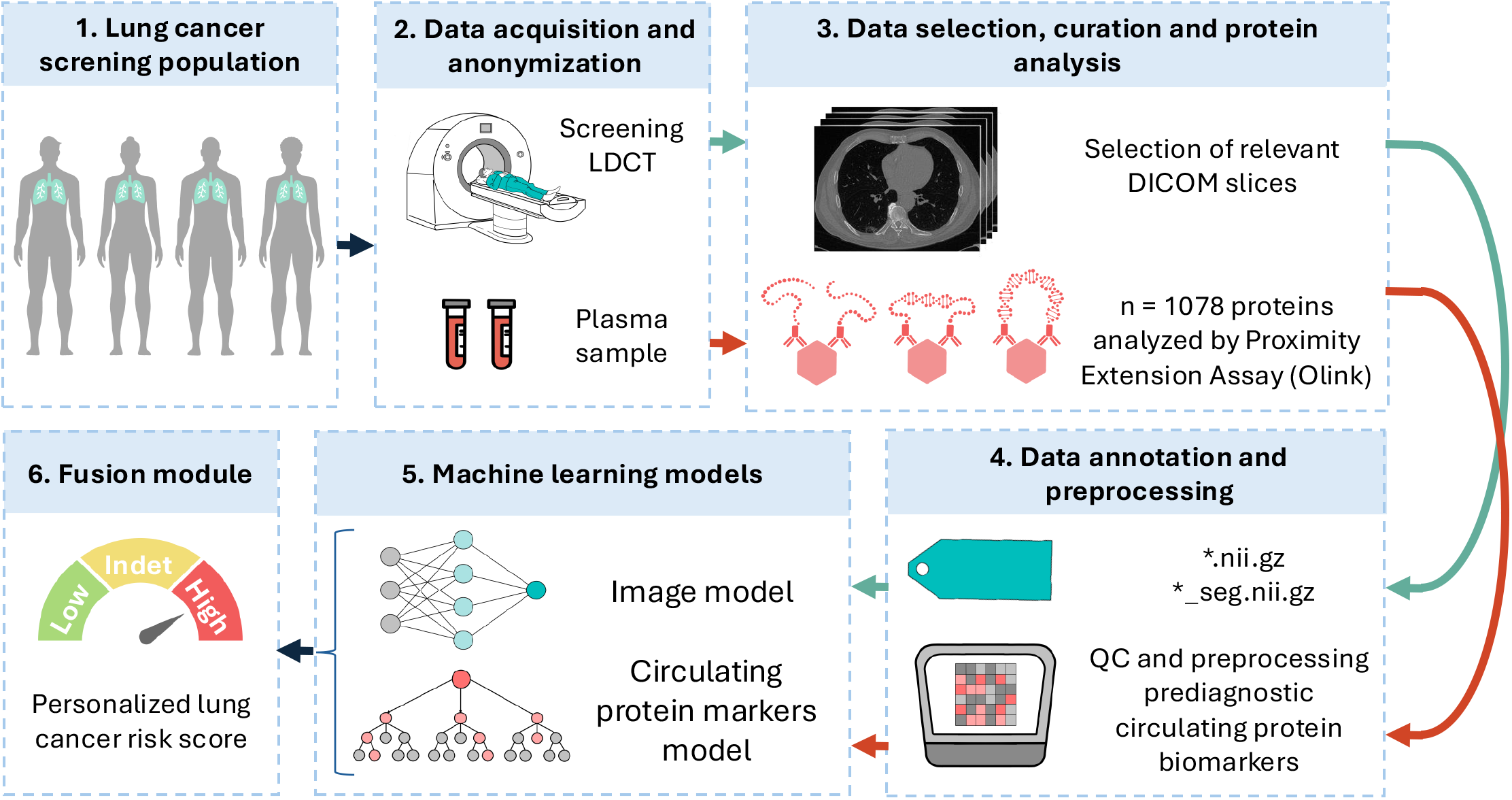
Workflow diagram. This diagram provides an overview of the dataset preparation and validation stages, including Lung cancer screening population, Data acquisition and anonymization, Data selection, curation and protein analysis, Data annotation, and preprocessing, Machine learning models and Fusion module.

### 2.1 Collection principles

To ensure precise subject selection and discard the impact of irrelevant factors, we adhered to the following principles in our case selection process:

#### Inclusion criteria

Participants were selected from a lung cancer screening population including men and women aged 40 years or older who were current or former smokers with a minimum smoking history of 10 pack-years, and who exhibited no symptoms of lung cancer at the time of enrollment. The specific inclusion criteria for P-ELCAP are further described in previous works by Sanchez-Salcedo *et al*. [34] and Mesa-Guzmán *et al*. [35]. Individuals with benign nodules were included as controls if the nodules remained stable in size during a follow-up period of at least two years.

#### Exclusion criteria

Participants with a prior history of lung cancer were excluded, as well as those diagnosed with any form of cancer within four years before or after enrollment, with the exception of non-melanoma skin cancer.

#### Pre-treatment imaging

LDCT scans were obtained before the administration of any relevant treatments and surgery, ensuring that the nodule’s imaging features were not altered by prior medical interventions.

#### Quality control

We conducted a comprehensive evaluation of LDCT images, discarding those with missing or incorrect layering to preserve the integrity and completeness of the lung nodule dataset.

### 2.2 Data collection and annotation

This cohort is part of the International Early Lung Cancer Action Program (P-ELCAP) conducted at Clínica Universidad de Navarra, Spain. This specific P-ELCAP subcohort was retrospectively collected and was also included in the Integrative Analysis of Lung Cancer Risk and Etiology (INTEGRAL) research program. Ethical approval was obtained from the University of Navarra Research Ethics Board (ref 2020.251, January 25th 2021). Informed consent was obtained from each participant individual. The dataset includes a total of 67 lung cancer cases collected within 5 years prior to diagnosis, 68 matched controls with benign pulmonary nodules, and 71 matched controls without lung nodules. For each case, sex, age and plasma sample collection time with respect to LDCT imaging were used to match control benign nodules and controls without nodules within 5 years. Lung cancer cases were considered if biopsy-confirmed lung cancer was diagnosed within 5 years. Additionally, there are 5 imaging false positives participants, who correspond to patients who underwent surgery to remove a suspicious nodule and, following resection, lung cancer was conclusively excluded by the pathologist as a diagnosis. The information available for each case was the output from proteomics analysis by 12 Olink Target 96 panels (more than 1000 proteins; see next section), as well as LDCT imaging data, together with clinical and demographic tabular data.

### 2.3 Data records and code

We have released the fully anonymized dataset, containing the aforementioned information for each case, in the EU-accredited repository Zenodo [36]. The dataset is deposited under a Creative Commons CC-BY-NC-SA license. This license enables users to distribute, remix, adapt, and build upon the material in any medium or format for noncommercial purposes only, and provided proper attribution is given to the creator both for the material or its modifications. The latest version of the informed consent and patient information datasheet was approved by the University of Navarra Research Ethics Committee on its June 2nd 2022 session. In the informed consent the screening participants provided permission for their data to be used exclusively in lung cancer research. As we do not foresee any applications beyond lung cancer, it is our view that all prospective users fall within the intended scope. Hence, a limitation in the Zenodo web access for lung cancer research is implemented just through a requester intention confirming box, and no formal Declaration of Use is requested. Thus, users will be granted access to the dataset in Zenodo^16^ upon stating through the request process that the dataset will be used exclusively for lung cancer research. The dataset’s file structure is shown in Fig 2, the different data modalities can be matched using the participant’s anonymized ID in the.csv and.xlsx files.

**Figure 2.**
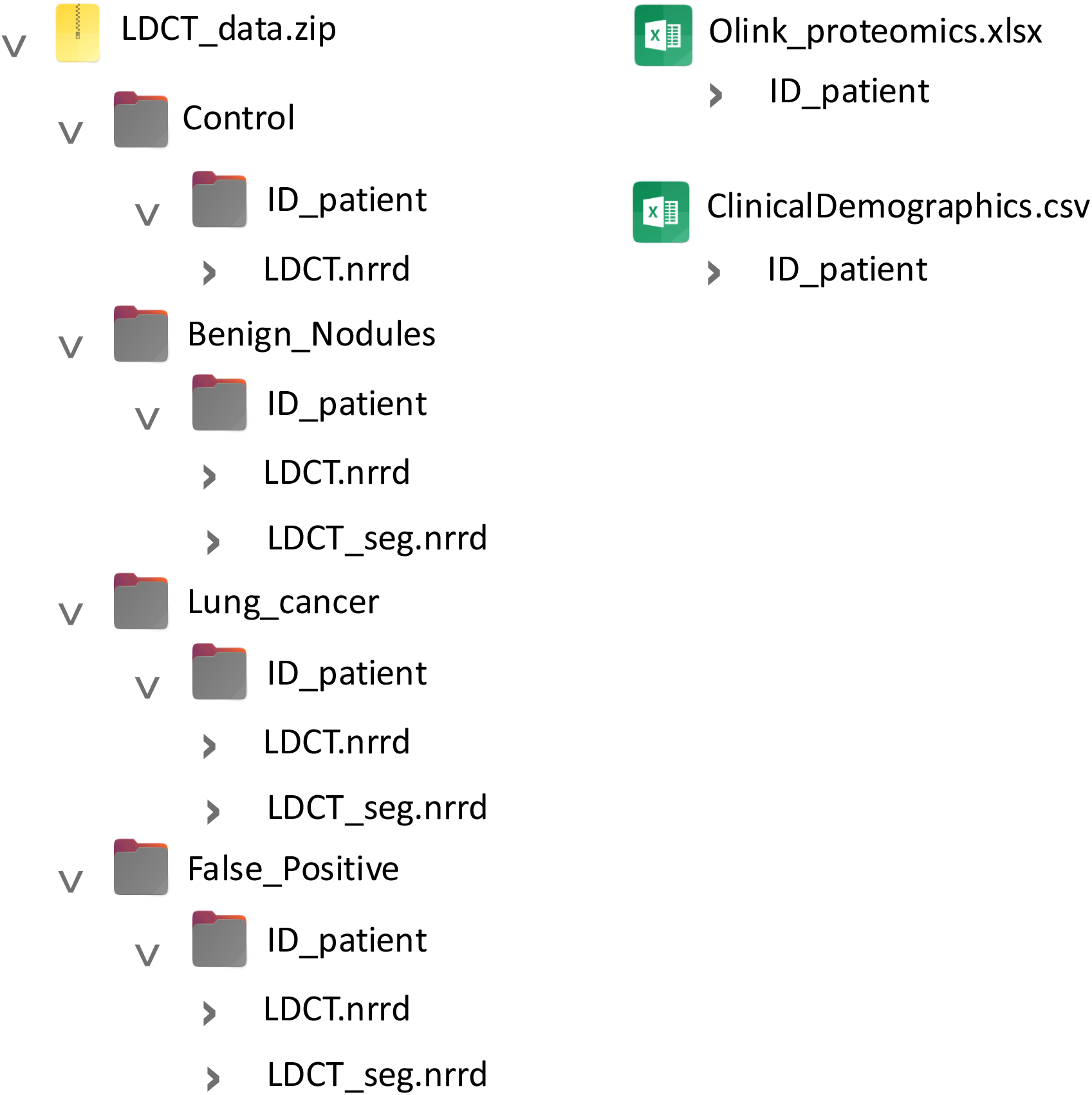
Structure and hierarchy of dataset files.

To ensure easy access to the code to read and process this dataset, we have made it available in a GitHub repository https://github.com/MiriamCobo/P-ELCAP_Dataset.git. The repository provides Python sample code demonstrating how to access and utilize the data, facilitating understanding and reuse.

## 3 Results

### 3.1 Dataset properties

Most participants were men (171 men vs. 40 women) and the median age at blood collection was 61 years (IQR 13 years). The median time between pre-diagnostic blood collection and diagnosis was 1.4 years (IQR 3 years, range: 0-5 years, by design). Of the participants diagnosed with lung cancer, 31 cases occurred at baseline or within the first year (using the date of blood and image collection as the reference baseline point). The remaining diagnoses occurred over subsequent years: 8 after one year, 9 after two years, 10 after three years, 7 after four years, and 2 after five years. Details on P-ELCAP cohort population characteristics are included in Table 1. The different data modalities included in the dataset are comprehensively described in the following paragraphs.

**Table 1:** Key characteristics of 67 lung cancer cases, 68 matched controls with benign pulmonary nodules, 71 matched controls without lung nodules and 5 false positives from P-ELCAP cohort. ^*a*^ N/A indicates that the patient is still alive; ^*b*^ Unknown indicates that there was no data available. Age was measured at the time of blood plasma collection.

| Characteristic | Cancer | Benign nodules | Controls | False positives | Total |
| --- | --- | --- | --- | --- | --- |
| **Sex n (%)** |  |  |  |  |  |
| Female | 12 (18.5%) | 13 (19.1%) | 13 (18.3%) | 2 (40.0%) | 40 (19.1%) |
| Male | 55 (82.1%) | 55 (80.9%) | 58 (81.7%) | 3 (60.0%) | 171 (81.0%) |
| **Age (years)** |  |  |  |  |  |
| Median (IQR) | 61.0 (14.5) | 61.5 (14.0) | 60.0 (12.0) | 68.0 (17.0) | 61.0 (13.0) |
| **Follow-up time (years)** |  |  |  |  |  |
| Median (IQR) | 8.0 (7.0) | 7.5 (6.0) | 7.0 (7.5) | 13.0 (7.0) | 7.0 (6.5) |
| **Follow-up survival time (years)** |  |  |  |  |  |
| Median (IQR) | 2.0 (4.0) | - | - | - | 2.0 (4.0) |
| N/A<sup>a</sup> n (%) | 46 (68.7%) | 68 (100%) | 71 (100%) | 5 (100%) | 190 (90.0%) |
| **Smoking status n (%)** |  |  |  |  |  |
| Current | 34 (50.7%) | 31 (45.6%) | 42 (59.2%) | 2 (40.0%) | 109 (51.7%) |
| Former | 33 (49.3%) | 37 (54.4%) | 29 (40.8%) | 3 (60.0%) | 102 (48.3%) |
| **Smoking duration (years)** |  |  |  |  |  |
| Median (IQR) | 41.3 (14.4) | 40.3 (15.2) | 38.6 (15.7) | 38.0 (10.0) | 40.4 (15.0) |
| **Smoking pack years** |  |  |  |  |  |
| Median (IQR) | 41.0 (36.1) | 31.8 (28.1) | 26.8 (22.4) | 42.0 (14.4) | 35.4 (28.0) |
| **Smoking quit years (former)** |  |  |  |  |  |
| Median (IQR) | 8.0 (11.8) | 8.9 (16.3) | 15.2 (19.1) | 12.7 (2.1) | 9.7 (15.0) |
| **Emphysema n (%)** |  |  |  |  |  |
| Yes | 30 (44.8%) | 15 (22.1%) | 15 (21.1%) | 0 (0.0%) | 60 (28.4%) |
| No | 37 (55.2%) | 53 (77.9%) | 56 (78.9%) | 5 (100.0%) | 151 (71.6%) |
| **COPD n (%)** |  |  |  |  |  |
| Yes | 41 (61.2%) | 23 (35.4%) | 23 (37.7%) | 2 (40.0%) | 89 (42.2%) |
| No | 26 (38.8%) | 42 (64.6%) | 38 (62.3%) | 3 (60.0%) | 109 (51.7%) |
| Unknown<sup>b</sup> | - | 3 (4.4%) | 10 (14.1%) | - | 13 (6.2%) |
| **Body Mass Index** |  |  |  |  |  |
| Median (IQR) | 26.2 (6.4) | 27.7 (5.1) | 27.3 (4.5) | 23.5 (4.0) | 26.9 (5.1) |
| **Family history of lung cancer n (%)** |  |  |  |  |  |
| Yes | 17 (25.4%) | 9 (13.2%) | 10 (14.1%) | 0 (0.0%) | 36 (17.1%) |
| No | 50 (74.6%) | 59 (86.8%) | 61 (85.9%) | 5 (100.0%) | 175 (82.9%) |
| **Personal history of cancer n (%)** |  |  |  |  |  |
| Yes | 4 (6.0%) | 8 (12.1%) | 5 (7.0%) | 3 (60.0%) | 20 (9.5%) |
| No | 63 (94.0%) | 58 (87.9%) | 66 (93.0%) | 2 (40.0%) | 189 (89.6%) |
| Unknown<sup>b</sup> | - | 2 (2.9%) | - | - | 2 (0.9%) |
| **Stage n (%)** |  |  |  |  |  |
| Early stage (TNM 1/2) | 53 (79.1%) | - | - | - | - |
| Advanced stage (TNM 3/4) | 14 (20.9%) | - | - | - | - |
| **Histology n (%)** |  |  |  |  |  |
| Adenocarcinoma | 38 (56.7%) | - | - | - | - |
| Squamous cell carcinoma | 12 (17.9%) | - | - | - | - |
| Large cell carcinoma | 9 (13.4%) | - | - | - | - |
| Small cell carcinoma | 4 (6.0%) | - | - | - | - |
| Mixed adenocarcinoma and squamous cell carcinoma | 2 (3.0%) | - | - | - | - |
| Other/NOS | 2 (3.0%) | - | - | - | - |

#### Olink proteomics

Relative protein concentrations of up to 1078 individual proteins across 12 panels focused on a specific area of disease or biology (Cardiometabolic, Cardiovascular II, Cardiovascular III, Development, Immune Response, Inflammation, Metabolism, Neurology, Oncology II, Oncology III, Organ Damage, NeuroExploratory) in prediagnostic plasma samples were measured with Olink Target 96 platform. Quantification measurement was carried out by Olink in their central laboratories in Uppsala, Sweden [31]. This high-throughput technology is based on the highly sensitive PEA technique, as previously described by the INTEGRAL consortium [31, 33]. Relative protein concentrations are reported as normalized protein expression (NPX) values on a log2 scale. These values are derived from quantitative PCR cycle threshold measurements and were standardized for subsequent analysis [31]. A detailed description of Olink methodology can be found in the white paper on Olink’s site [37].

#### LDCT scans

For each LDCT, we selected the series with the thinnest CT image slices, defined as having a slice thickness of ≤ 3 mm, for inclusion in the dataset. LDCT scans were collected within a 6-month window (184 days), either before or after the date of blood plasma collection. For 5 lung cancer cases the nodule was not visible in the LDCT scan at the time of the plasma sample. Lung nodules were annotated by an experienced radio-technician and subsequently supervised, refined, and approved by a certified experienced thoracic radiologist, who is a member of the P-ELCAP LDCT multidisciplinary team. Figure 3 shows horizontal bar plots of nodule dimensions (width, height, and depth), and the anatomical location of the nodules. We additionally show one example of LDCT scans for a participant belonging to each of the groups in Figure 4.

**Figure 3.**
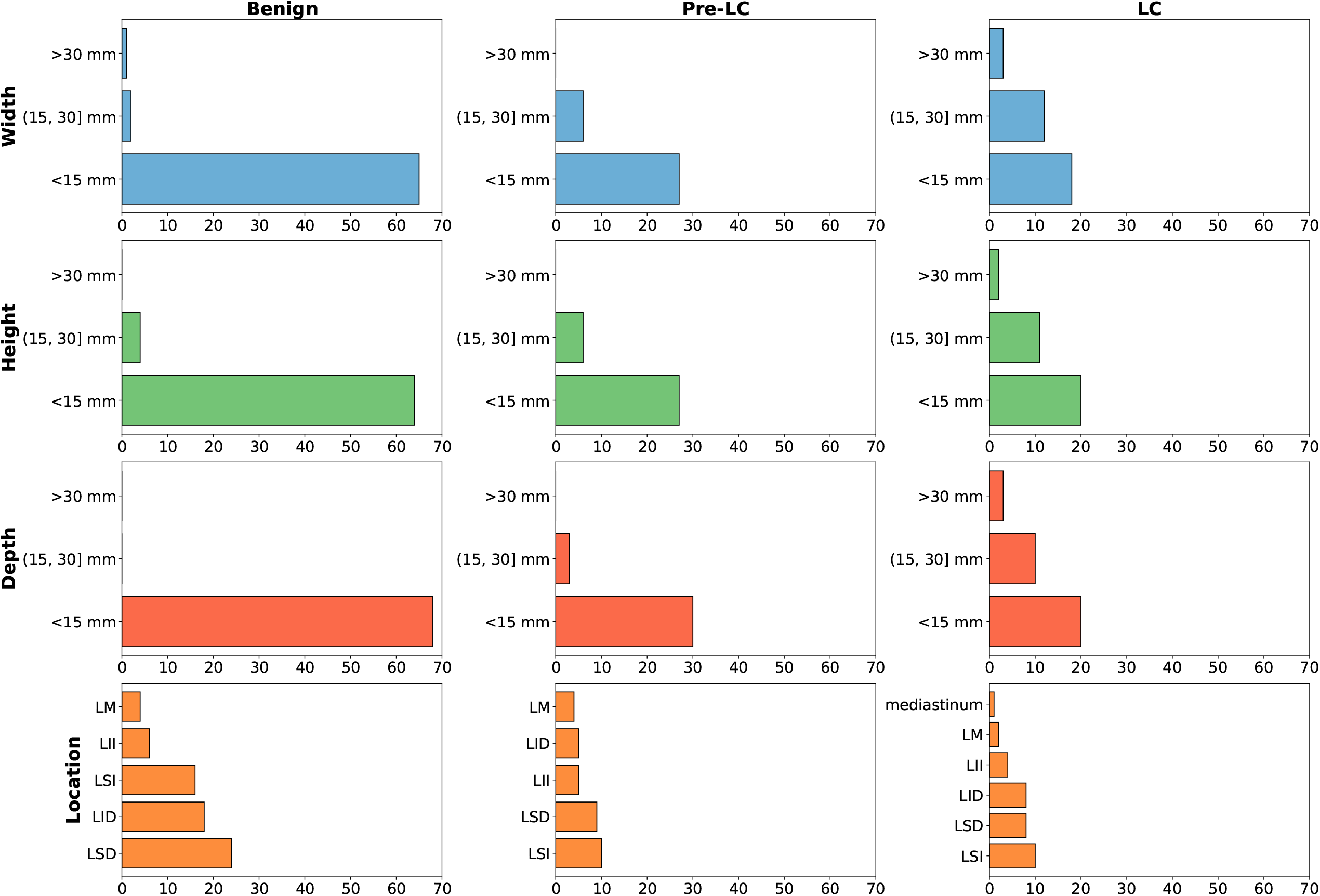
Distribution of lung nodule sizes and locations across diagnostic groups. The x-axis denotes the nodule count. These groups include controls with benign nodules (“Benign”), lung cancer from 1-5 years before diagnosis (“Pre-LC”), and lung cancer less than 1 year before diagnosis (“LC”).

**Figure 4.**
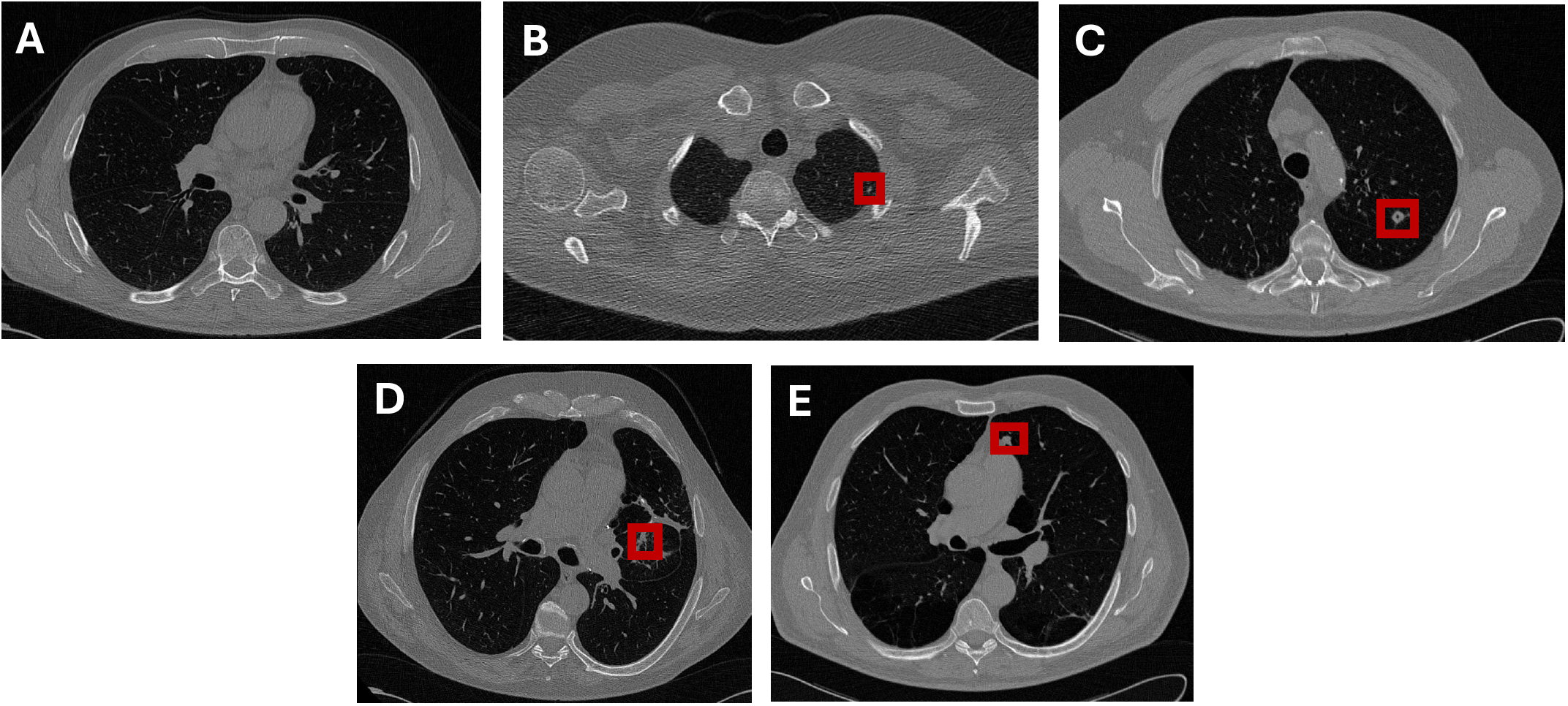
Example of a LDCT image. **A:** Control without nodules. **B:** Control with benign nodule. **C:** Imaging false positive. **D:** Lung cancer from 1-5 years before diagnosis. **E:** Lung cancer less than 1 year before diagnosis.

#### Clinical and demographic data

The clinicians involved in the P-ELCAP multidisciplinary team (mainly neumologists and specialized nurses) collected the demographic/clinical information of the study participants (age, sex, smoking questionnaire, comorbidities, exposures, lifestyle, etc.). The basic information (age, sex, smoking) is included in the available public information for each individual in the Zenodo dataset (see below). All cases included in the study are of Caucasian ethnicity. The participant age in the dataset corresponds to the date of blood collection. We also include in the open access dataset the difference in years and in months between the date of blood collection and the date of diagnosis, rounding to the nearest month unit. For control participants (with or without nodules), this time was set to the maximum follow-up time observed among lung cancer participants (5 years or 60 months) to enable the use of time-to-event models such as random survival forest. We include the following variables: Group, Age, Sex, Stage, Smoking (in pack years), and the aforementioned time in years and months.

### 3.2 Technical evaluation

To validate the dataset proposed in this study, we have performed a proof-of-concept/technical validation study using the data available in the open access dataset. The aim is to conduct two baseline studies for technical validation: image-based deep learning (DL) models from the available LDCT images; and machine learning (ML) protein models from Olink proteomics data. Data was divided into 3 fold cross-validation (CV) train and test splits, and in the image models a validation subset representing 10% of training data was separated. Data was stratified according to Figure 5. The same 3 folds were used to train and evaluate all the methods, filtering by the groups of participants depending on the case. All the analyses were done in Python. Performance was evaluated using binary classification metrics: area under the curve (AUC), accuracy, balanced accuracy, precision (i.e., positive predictive value, PPV), recall (i.e., sensitivity), F1-score, specificity and negative predictive value (NPV). Unless otherwise specified, a threshold of 0.5 was used to compute the binary values. In the following sub-sections we describe in more detail the two proof-of-concept studies.

**Figure 5.**
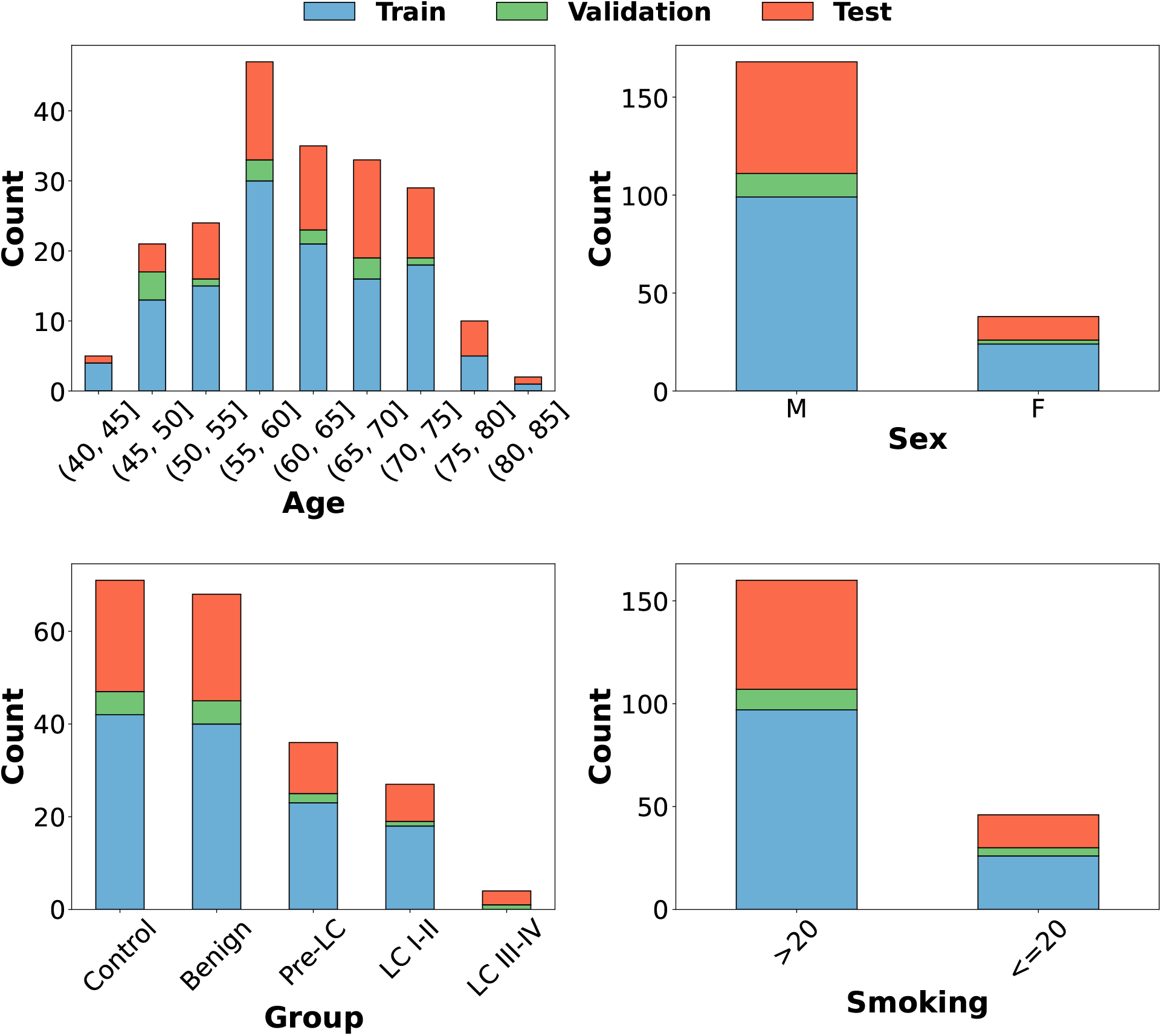
Stratification of data splits (train, validation, test) in the first fold according to age, sex, group and smoking (in pack years) variables. In the panel labeled “Group”, “Control” corresponds to controls without nodules, “Benign” to controls with nodules, “Pre-LC” to lung cancer cases from 1 to 5 years prior diagnosis, “LC I-II” to lung cancer cases at stages I-II, and “LC III-IV” to lung cancer cases at stages III-IV.

#### 3.2.1 Image models

LDCT images from control participants with benign nodules and the corresponding lung cancer participants diagnosed in the same year as the blood collection were included in this analysis. We explored the effectiveness of 3D DL architectures for lung nodule classification. A crop around the nodule’s centroid, determined with the segmentation, with 3D size [32, 32, 32] in pixels was performed to include surrounding information. Nodules larger than the specified size in any dimension were down-sampled in that dimension to ensure compatibility with the network’s architecture. We developed the models (DenseNet121, EfficientNet, ResNet34) using MONAI [38] and Pytorch, incorporating pretrained weights from Med3D when available to enhance performance [39]. A weighting strategy was used in the loss function to improve learning from imbalanced class distributions. We included small data augmentations in the train subset with a probability of 50% (random rotations in a range of 30° and isotropic scaling by a factor uniformly sampled between 0.9 and 1.1.), and dropout to enhance the learning of the model [40] and improve generalization. Results are shown in Table 2. The results are promising in some of the comparative indicators (AUC, accuracy, PPV, NPV) especially in the EfficientNet analysis, but at the same time reveal the limitations of training deep architectures with a small dataset size [40]. What is clear is the potential of this open dataset if used in combination with other datasets, which eventually may become available in the future by other groups conducting research on lung cancer screening cohorts.

**Table 2:** Results of the deep learning models trained on LDCT data for predicting benign participants and patients diagnosed with lung cancer in the same year as the plasma collection, evaluated using 3-fold cross-validation test sets. All models were implemented in their 3D architecture.

| Metric | DenseNet121 | EfficientNet | ResNet34 |
| --- | --- | --- | --- |
| Pretraining | No | No | Yes |
| AUC | 0.50 ± 0.07 | 0.86 ± 0.04 | 0.66 ± 0.05 |
| Accuracy | 0.43 ± 0.06 | 0.80 ± 0.04 | 0.60 ± 0.08 |
| Balanced Accuracy | 0.51 ± 0.10 | 0.76 ± 0.09 | 0.61 ± 0.09 |
| Precision/PPV | 0.33 ± 0.07 | 0.74 ± 0.05 | 0.3 ± 0.3 |
| Recall/Sensitivity | 0.76 ± 0.19 | 0.6 ± 0.2 | 0.6 ± 0.5 |
| F1-score | 0.46 ± 0.09 | 0.66 ± 0.14 | 0.4 ± 0.3 |
| Specificity | 0.26 ± 0.01 | 0.88 ± 0.07 | 0.6 ± 0.4 |
| NPV | 0.72 ± 0.16 | 0.84 ± 0.08 | 0.84 ± 0.16 |

#### 3.2.2 Protein models

Almost 1000 circulating proteins were used in this second part of the proof-of-concept study. Markers with missing values or that failed to reach Olink’s quality control were excluded from our technical validation; however, in the open database available at Zenodo, the raw values of these proteins are also provided as they may be useful to support some other type of analyses, allowing for missing values. This filtering resulted in a total of 970 proteins. The same 3 fold CV previously described with training and test subsets was used. The NPX (Normalized Protein eXpression) data (see OLINK whitepaper [37]) were processed using standard scaler trained on the training subset (comprising both the training and validation sets), and then subsequently applied to the test subset. The best model for each fold was found through another grid search 3 fold CV on the training subset, and then it was evaluated on the separated test subset of that fold. We trained three ML models: random forest (RF), xgboost (XGB) and penalized regression (LASSO) to distinguish benign versus lung cancer participants diagnosed in the same year as plasma collection. We additionally trained a novel architecture on biologically informed neural networks (BINN) [41], that leverages the pathways from Reactome pathway database [42] to create a sparse neural network. Results are shown in Table 3. Furthermore, a random survival forest (RSF) was trained on all lung cancer and benign participants, oversampling lung cancer participants in the training set, that were repeated. Results for participants in years 0, 1, and 2 previous to lung cancer diagnosis are reported in Table 4. The most relevant proteins identified by the models are listed in Table 5. These proteins were calculated first identifying the 100 most relevant proteins in each fold, and subsequently those that were repeated across the 3 folds were included in the list.

**Table 3:** Results of the machine learning models trained on protein data for predicting benign participants and patients diagnosed with lung cancer in the same year as the plasma collection, evaluated using 3-fold cross-validation test sets.

| Metric | RF | XGB | LASSO | BINN |
| --- | --- | --- | --- | --- |
| AUC | 0.65 ± 0.08 | 0.65 ± 0.03 | 0.71 ± 0.07 | 0.59 ± 0.19 |
| Accuracy | 0.72 ± 0.08 | 0.71 ± 0.06 | 0.71 ± 0.05 | 0.57 ± 0.13 |
| Balanced Accuracy | 0.61 ± 0.05 | 0.63 ± 0.01 | 0.62 ± 0.08 | 0.57 ± 0.16 |
| Precision/PPV | 0.7 ± 0.3 | 0.58 ± 0.14 | 0.52 ± 0.11 | 0.38 ± 0.16 |
| Recall/Sensitivity | 0.32 ± 0.05 | 0.41 ± 0.07 | 0.39 ± 0.19 | 0.57 ± 0.20 |
| F1-score | 0.42 ± 0.07 | 0.47 ± 0.01 | 0.44 ± 0.17 | 0.45 ± 0.18 |
| Specificity | 0.90 ± 0.09 | 0.84 ± 0.10 | 0.85 ± 0.02 | 0.57 ± 0.10 |
| NPV | 0.74 ± 0.04 | 0.76 ± 0.02 | 0.76 ± 0.06 | 0.75 ± 0.11 |

**Table 4:**
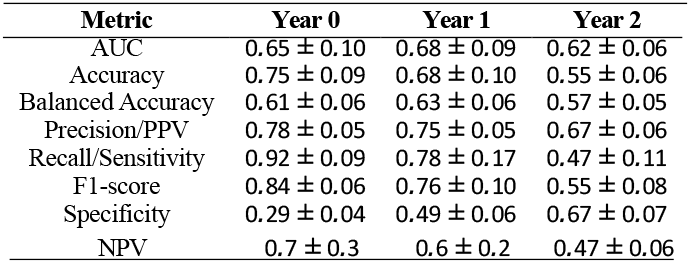
Results of the random survival forest trained on protein data for predicting benign participants and patients diagnosed with lung cancer at different time points, evaluated using 3-fold cross-validation test sets. The classification threshold was empirically set at 0.6, as this value provided a better trade-off between sensitivity and specificity across the evaluated folds.

| Metric | Year 0 | Year 1 | Year 2 |
| --- | --- | --- | --- |
| AUC | 0.65 ± 0.10 | 0.68 ± 0.09 | 0.62 ± 0.06 |
| Accuracy | 0.75 ± 0.09 | 0.68 ± 0.10 | 0.55 ± 0.06 |
| Balanced Accuracy | 0.61 ± 0.06 | 0.63 ± 0.06 | 0.57 ± 0.05 |
| Precision/PPV | 0.78 ± 0.05 | 0.75 ± 0.05 | 0.67 ± 0.06 |
| Recall/Sensitivity | 0.92 ± 0.09 | 0.78 ± 0.17 | 0.47 ± 0.11 |
| F1-score | 0.84 ± 0.06 | 0.76 ± 0.10 | 0.55 ± 0.08 |
| Specificity | 0.29 ± 0.04 | 0.49 ± 0.06 | 0.67 ± 0.07 |
| NPV | 0.7 ± 0.3 | 0.6 ± 0.2 | 0.47 ± 0.06 |

**Table 5:**
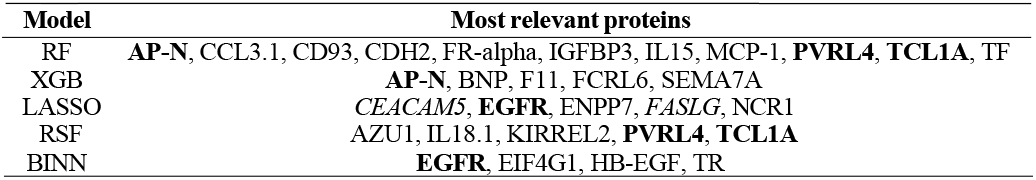
Proteins ranked among the top 100 in predicting benign participants and patients diagnosed with lung cancer in the same year as the plasma collection across all three folds based on feature importance analysis. In **bold**, proteins that are repeated across different methods. In *italic*, proteins that were in the list of the 10 protein markers most relevant for imminent lung cancer in a previous work within the INTEGRAL consortium [33].

| Model | Most relevant proteins |
| --- | --- |
| RF | **AP-N**, CCL3.1, CD93, CDH2, FR-alpha, IGFBP3, IL15, MCP-1, **PVRL4**, **TCL1A**, TF |
| XGB | **AP-N**, BNP, F11, FCRL6, SEMA7A |
| LASSO | *CEACAM5*, **EGFR**, ENPP7, *FASLG*, NCR1 |
| RSF | AZU1, IL18.1, KIRREL2, **PVRL4**, **TCL1A** |
| BINN | **EGFR**, EIF4G1, HB-EGF, TR |

## 4 Discussion

P-ELCAP dataset is a valuable lung cancer screening cohort to support open access research. This dataset comprises a small-medium size cohort including 67 lung cancer patients, 68 controls with benign pulmonary nodules, 71 controls without nodules and 5 imaging false positives. To the best of our knowledge, this is the first multimodal open access database including long-term follow-up benign and biopsy confirmed malignant pulmonary nodules, as well as controls without nodules. Furthermore, this is the first publicly available lung cancer screening dataset incorporating over 1000 circulating protein markers generated by PEA. The applications of this dataset for lung cancer research extend beyond the scope of this work, which focuses on assessing data quality and leaves further discoveries to future investigations. The technical validation results underscore the potential of the data, as well as inherent challenges associated with the limited dataset size. The LDCT image-based models can be further refined leveraging other publicly available datasets to enhance current results, that as a proof-of-concept were only trained on P-ELCAP dataset. The proposed protein models identified a set of proteins that consistently appear across different methods, demonstrating robustness and alignment with findings from previous studies [33]. Moreover, the availability of a set of around 1000 circulating proteins in individuals with malignant versus benign nodules may open a wealth of research hypothesis or help as a validation tool for discovery projects performed in similar or even distant clinical settings. The field of characterization of incidental nodules may highly benefit from the data made available in the present database.

P-ELCAP dataset holds significant value for external validation in other screening cohorts, and represents an important step toward advancing the field of personalized medicine through AI-driven multimodal models in the context of LDCT-based lung cancer screening. Our dataset will also help in the development and validation of novel multimodal strategies in early lung cancer screening. Future work may enhance the proposed image models through pretraining in larger publicly available LDCT imaging datasets, with evaluation on our LDCT cohort to investigate the generalizability of existing DL approaches for multimodal lung nodule detection and risk characterization. Although we provide a highly useful tool for research in this field, further work and external validation with other cohorts is necessary to reach real life clinical utility of multimodal tools in LDCT-based lung cancer screening.

## Data Availability

All data produced in the present study are available upon reasonable request to the authors

https://doi.org/10.5281/zenodo.15120062

## Acknowledgments

M. C. would like to acknowledge the support received by the Ministry of Education of Spain (FPU grant, reference FPU21-04458). M.C. And L.L.I. would like to acknowledge the support from the project AI4EOSC “Artificial Intelligence for the European Open Science Cloud” that has received funding from the European Union’s Horizon Europe research and innovation programme under grant agreement number 101058593. L. M. M. and G. B. received a Lung Ambition Alliance award grant for this project. L. M. M. was also supported by CIBERONC (CB16/12/00443), Spanish Ministry of Science and Innovation and Fondo de Investigación Sanitaria Fondo Europeo de Desarrollo Regional (PI22/00451). D. O. was supported by by the Ministry of Education of Spain (FPU grant, reference FPU20/06292). D. S. is a recipient of a Ramón y Cajal grant (# RYC2022-038084-I) funded by MICIU/AEI/10.13039/501100011033 and by “ESF+”.

## Ethics statement

The authors declare that this manuscript represents entirely original works, and/or if work and/or works of others have been used, that this has been appropriately cited or quoted. This material has not been published in whole or in part elsewhere. The manuscript is not currently being considered for publication in another journal. The main aim of the present manuscript is to promote Open Science collaborations in our research area.

## Author contributions

Conceptualization: M.C., L.L.I., D.S., L.M.M. Methodology: M.C., L.L.I., D.S., L.M.M. Software: M.C. Validation: M.C., L.L.I., D.S., L.M.M. Formal analysis: M.C. Investigation: M.C. Resources: D.S., J.B., A.P., J.P.T., J.J.Z., J.I.E., A.E., J.P., A.A., J.S.O., J.B., A.B.A., M.D.F., C.F., A.C., A.F., A.E., K.V., D.O., M.M.O., H.B.G., M.A.F., V.P., M.R., M.D.L., A.C., R.P., R.J.H., L.M-S., G.B., L.L.I., L.M.M. Data curation: M.C, L.L.I., D.S., J.B., A.P., G.B., L.M.M. Writing - Original Draft: M.C. Writing - Review & Editing: M.C., L.L.I., D.S., L.M.M, A.C., W.S. Visualization: M.C. Supervision: L.L.I., L.M.M, G.B., L.M.S. Project administration: L.L.I., L.M.M, R.J.H., L.M.S., G.B. Funding acquisition: L.M.M.

## Authors statement

All authors have read and approved the manuscript, and it has not been accepted for publication

## Footnotes

16 For reviewers, a representative sample of the dataset has been uploaded to https://nextcloud.ifca.es/index.php/s/aWJGHTfkTQgBTeg

